# Whole-Genome Sequencing Reveals Individual and Cohort Level Insights into Chromosome 9p Syndromes

**DOI:** 10.1101/2025.03.28.25324850

**Authors:** Yingxi Wang, Eleanor I. Sams, Rachel Slaugh, Sandra Crocker, Emily Cordova Hurtado, Sophia Tracy, Ying-Chen Claire Hou, Christopher Markovic, Kostandin Valle, Victoria Tate, Khadija Belhassan, Elizabeth Appelbaum, Titilope Akinwe, Rodrigo Starosta Tzovenos, Yang Cao, Amber Neilson, Yu Liu, Nathaniel Jensen, Reza Ghasemi, Tina Lindsay, Juana Manuel, Sophia Couteranis, Milinn Kremitzki, Jack Ustanik, Thomas Antonacci, Jeffrey K. Ng, Andrew Emory, Laura Metz, Tracie DeLuca, Katherine N. Lyons, Toni Sinnwell, Brianne Thomeczek, Kymme Wang, Nick Sisneros, Megha Muraleedharan, Anantha Kethireddy, Marco Corbo, Harsha Gowda, Katherine King, Christina A. Gurnett, Susan K. Dutcher, Catherine Gooch, Yang E. Li, Matthew W. Mitchell, Kevin A. Peterson, Amjad Horani, Jill A. Rosenfeld, Weimin Bi, Pawel Stankiewicz, Hsiao-Tuan Chao, Jennifer Posey, Christopher M. Grochowski, Zain Dardas, Erik Puffenberger, Christopher E. Pearson, Frank Kooy, Dale Annear, A. Micheil Innes, Michael Heinz, Richard Head, Robert Fulton, Stephan Toutain, 9P-ARCH, Lucinda Antonacci-Fulton, Xiaoxia Cui, Robi D. Mitra, F. Sessions Cole, Julie Neidich, Patricia I. Dickson, Jeffrey Milbrandt, Tychele N. Turner

**Author notes:** co-first authors (Yingxi Wang, Eleanor I. Sams, Rachel Slaugh, Sandra Crocker). senior authors (Julie Neidich, Patricia I. Dickson, Jeffrey Milbrandt, Tychele N. Turner). Correspondence to: Tychele N. Turner, Ph.D.

## Abstract

Previous genomic efforts on chromosome 9p deletion and duplication syndromes have utilized low resolution strategies (i.e., karyotypes, chromosome microarrays). We present the first large-scale whole-genome sequencing (WGS) study of 100 individuals from families with 9p-related syndromes including 85 unrelated probands through the 9P-ARCH (**A**dvanced **R**esearch in **C**hromosomal **H**ealth: Genomic, Phenotypic, and Functional Aspects of **9p**-Related syndromes) research network. We analyzed the genomic architecture of these syndromes, highlighting fundamental features and their commonalities and differences across individuals. This work includes a machine-learning model that predicts 9p deletion syndrome from gene copy number estimates using WGS data. Two Late Replicating Regions (LRR1 [a previously un-named human fragile site], LRR2) were identified that contain most structural variant breakpoints in 9p deletion syndrome pointing to replication-based issues in structural variant formation. Furthermore, we show the utility of using WGS information to obtain a comprehensive understanding of 9p-related variation in an individual with complex structural variation where chromothripsis is the likely mechanism. Genes on 9p were prioritized based on statistical assessment of human genomic variation. Furthermore, through application of spatial transcriptomics to embryonic mouse tissue we examined 9p-gene expression in craniofacial and brain development. Through these strategies, we identified 24 important genes for the majority (83%) of individuals with 9p deletion syndrome including *AK3, BRD10, CD274, CDC37L1, DMRT1, DMRT2, DMRT3, DOCK8, GLIS3, JAK2, KANK1, KDM4C, PLPP6, PTPRD, PUM3, RANBP6, RCL1*, *RFX3*, *RIC1*, *SLC1A1*, *SMARCA2*, *UHRF2*, *VLDLR*, and *ZNG1A*. Two genes (*AK3*, *ZNG1A*) are involved in mitochondrial function and testing of the mitochondrial genome revealed excess copy number in individuals with 9p deletion syndrome. This study presents the most comprehensive genomic analysis of 9p-related syndromes to date, with plans for further expansion through our 9P-ARCH research network.

## INTRODUCTION

Syndromes involving the deletion or duplication of the short arm of chromosome 9 (9p) are characterized by loss and/or gain of DNA sequence on the p-arm of chromosome 9. These syndromes exhibit substantial phenotypic variability, with neurodevelopmental disorders (NDDs) being the most common feature across affected individuals ^1^. Most genetic and genomic studies on these syndromes focus on karyotypes or chromosome microarray data providing limited resolution to the structural variation. An international family network of individuals with 9p deletion syndrome now includes individuals with 9p duplications. In our previous work, we examined >700 individuals for their karyotypes and found that the most common deletions in 9p deletion syndrome encompass the region from the telomere through 9p24 or the region from the telomere to 9p22 ^1^. Few individuals have been identified with deletions near the centromere of 9p. In that study, which was the largest to look at genomic variation in individuals with this syndrome to date, we achieved “Precision Genomics” for one individual by using advanced genomic sequencing technologies ^1^. The variation within the individual was quite complex and set a precedent for the work in this current study. Recently, we examined phenotypes in 48 individuals with 9p deletion syndrome highlighting several shared physical characteristics (e.g., NDDs, dysmorphic features) ^2^ and advancing previous phenotype-related analyses in this area ^3–11^.

There are two main areas of investigation in the present study. The first is to analyze the genomes at the level of the individual and the second is cross-cohort analyses. At the level of the individual, a primary goal is to resolve the structural variation within individuals with these syndromes to the precise nucleotide so that the specific genes can be identified within the structural variation. Furthermore, since reports have suggested that 50% of individuals harbor a 9p deletion and a related translocation it is critical to identify the translocations since they may affect the expression of genes. Cross-cohort analyses are important for finding the genes that are most important across all individuals with this syndrome to lead to insights for diagnostics and therapeutics.

Several unique factors make this study extremely timely regarding providing key insights into these syndromes. We connected with the family-based Chromosome 9P Minus Network to pursue rigorous research studies of 9p-related syndromes. As part of this work, we also engaged the larger rare disease community to identify more individuals with these syndromes to form a 9p Research Network that we call **9P-ARCH** – **A**dvanced **R**esearch in **C**hromosomal **H**ealth: Genomic, Phenotypic, and Functional Aspects of **9p**-Related syndromes. This outreach allowed us to gain access to the largest 9p cohort (and family members) analyzed to date.

A unique aspect of our work is implementation of advanced genomic sequencing technologies with capabilities to perform short-read whole-genome sequencing on 100 individuals with 9p-related syndromes and their family members. Importantly, both the nuclear and mitochondrial genomes can be studied with this technology. Other genomic technologies are also useful to supplement the short-read whole-genome sequencing data and are pursued herein in more limited cases. By short-read whole-genome sequencing and sophisticated computational analyses of this data, we pursue the first detailed gene-level analyses on the p-arm of chromosome 9 in these syndromes. In this research, we also benchmark a machine-learning model to identify 9p-related syndromes from gene dosage alone.

Another aspect related to our work is the availability of large-scale genomic studies of individuals with neurodevelopmental disorders and individuals who have no obvious 9p-related phenotypes. In the present study, we leverage denovo-db ^12^, which now contains > 1 million *de novo* variants (DNVs) in ∼70,000 parent-child sequenced trios. We specifically focus on DNVs on 9p in 62,181 of these individuals with relevant phenotypes to provide further information regarding the importance of these genes through statistical enrichment models. We also analyze whole-genome sequencing data in 5,824 individuals with autism to examine potential enrichment of DNVs in noncoding, regulatory regions of the genome using existing and new statistical methods. In addition, we also examine large datasets of individuals without neurodevelopmental disorders which allows us to test dosage and constraint of genes ^13,14^. These datasets provide critical information about genes on 9p which we then directly apply to the analysis of genomes of individuals with 9p-related syndromes. Finally, through modern spatial expression techniques, we analyze the embryonic expression of the top 9p-related genes in the developing mouse to characterize different areas of the face and brain. These data provide new insights into gene activity and potential relevance to 9p-related syndromes. Overall, this work represents the most comprehensive genomic analysis of 9p-related syndromes to date.

## MATERIALS AND METHODS

### WashU 9p Deletion and Duplication Cohort

To establish a collection of individuals with 9p deletions and/or duplications we assembled a team of clinical, genomic and functional researchers. In collaboration with the Chromosome 9P Minus Network, which is a family network, we recruited 87 individuals from families with 9p deletion and/or duplication syndrome. Included were 70 unrelated probands, six fathers, seven mothers, one grandfather, one grandmother, and two affected siblings. Everyone was consented into the study using an age-appropriate form of consent/assent and in accordance with the Human Research Protection Office at Washington University in St. Louis (IRB #201706062). Blood samples were obtained for karyotype, DNA isolation for genomic analyses, and PBMC purification for downstream cell analyses.

All participants in the WashU 9p Deletion and Duplication Cohort underwent at least one of the following genomic assessments: clinical genomic record review, karyotype, Illumina short-read whole-genome sequencing, Bionano Optical Mapping, PacBio HiFi long-read whole-genome sequencing, SNP microarray, or Illumina CLR whole-genome sequencing. For the clinical genomic record review, the records were reviewed by a minimum of two individuals for derivation of variant details. Karyotype generation and assessment was performed at the Washington University in St. Louis Clinical Genomics/Cytogenetics and Molecular Pathology Core Lab. Cells were hypotonic treated, fixed, and then stained with GTG banding and examined with a microscope. Cells were analyzed using GTW banding method, and 20 metaphases were examined for each individual. The other genomic technologies were carried out at the Washington University in St. Louis McDonnell Genome Institute. Illumina short-read whole-genome sequencing was achieved to an average coverage of 30× using an Illumina Novaseq 6000 sequencer on a total of 84 individuals and is the predominant data source underlying the analyses in this study. Bionano Optical Mapping was fulfilled using a Bionano Saphyr system. PacBio HiFi long-read whole-genome sequencing was executed to an average coverage of 20× on a PacBio SequelII sequencer. SNP microarray was accomplished using a ThermoFisher CytoScan HD array. Illumina CLR whole-genome sequencing was completed to an average coverage of 30× using an Illumina Novaseq 6000 sequencer.

### Cohort of Individuals with 9p Deletion and/or Duplication Syndromes from NIGMS Repository

The NIGMS Human Genetic Cell Repository at the Coriell Institute for Medical Research (https://www.coriell.org/1/NIGMS/Collections/Chromosomal-Abnormalities) was queried for individuals with genomic variation on the p-arm of chromosome 9. Candidate samples were identified and checked for available cells. In total, there were 16 samples with available cells for performing DNA extraction. Both standard DNA extraction ^15^ and high molecular weight DNA extraction ^16^ was performed on these samples in the Coriell Institute’s Molecular Biology Laboratory. All 16 samples underwent Illumina short-read whole-genome sequencing to average coverage of 30× at the Washington University in St. Louis McDonnell Genome Institute. Additionally, eight of the samples were sequenced on a PacBio Revio sequencer to an average coverage of 30× at MedGenome.

### Aggregation of Additional Individuals with 9p Deletion and/or Duplication Syndromes

We contacted several genomics researchers across the United States of America to identify additional individuals with 9p Deletion and/or Duplication Syndromes and identified 26 individuals. These individuals had previously been assessed using SNP microarray technologies.

### Bioinformatic Assessment of Genomic Data

Illumina Short-Read Whole-Genome Sequencing: Post-sequencing, the fastq files were aligned to GRCh38_full_analysis_set_plus_decoy_hla.fa (build 38 of the human genome) using bwa mem ^17^ version 0.7.17-r1188, followed by sorting with SAMtools ^18^ version 1.9 sort, and indexing with SAMtools version 1.9 index. Coverage metrics were calculated using Picard ^19^ 2.25.4 CollectWgsMetrics. Structural variants were called using manta ^20^ version 1.6.0. Mitochondrial genome assessment was performed using EGP ^21^ version 1.3. Windowed copy number estimates across the genome were generated using QuicK-mer2 ^22^. SNV/indel calls were generated using DeepVariant ^23^ version 1.0.0. STRs were detected using ExpansionHunter ^24^ and GangSTR ^25^. Copy number genotyping was performed using CNPI (https://github.com/tnturnerLab/cnpi). For parent-child sequenced trios, DNVs were detected with HAT^26,27^ and quality checked with acorn ^28^.

PacBio Long-Read Whole-Genome Sequencing: Post-sequencing either the bam or fastq files were either submitted to the primrose program or directly aligned to GRCh38_full_analysis_set_plus_decoy_hla.fa (build 38 of the human genome) using pbmm2 version 1.10.0 followed by sorting with SAMtools ^18^ version 1.9 sort, and indexing with SAMtools version 1.9 index. Copy number assessment was performed using hificnv. SNV/indel calls were generated using DeepVariant ^23^ version 1.0.0.

Bionano Optical Mapping was analyzed using the Bionano Genomics software platform, SNP microarray was analyzed using ThermoFisher CytoScan HD Suite, and Illumina CLR whole-genome sequencing was analyzed in Illumina Dragen.

### Dosage of Genes on 9p

Per-gene copy number estimates were generated from whole-genome sequencing data from the 100 individuals with 9p-related syndromes and their family members in this study and 2,504 unrelated individuals in the 1000 Genomes Project ^29^. The individuals with 9p deletion syndrome and the individuals from the 1000 Genomes Project were compared gene-by-gene for differences in copy number using a Mann-Whitney U test. This data was also used in two distinct machine-learning model strategies including Random Forest and a Naïve Bayes. R packages used in the machine-learning analyses included caret ^30^, e1071 (https://CRAN.R-project.org/package=e1071), randomforest (https://cran.r-project.org/web/packages/randomForest). For machine learning, the 9p deletion syndrome individuals consisted of the 68 individuals that had deletions on 9p, in our analyses, and the control individuals consisted of the 2,504 unrelated individuals from the 1000 Genomes Project. For each of these datasets, 70% of the data was utilized in the training and 30% of the data was used for the testing. Several metrics were calculated to benchmark the models including accuracy, sensitivity, specificity, positive prediction value, and negative prediction value.

### Updated denovo-db

For this study, we updated denovo-db ^31^ to version 1.8 which is a database of DNVs aggregated from the published literature. In this version, there are 1,131,762 DNVs from 72,794 parent-child sequenced trios. These DNVs were compiled from 81 studies and include data on the following phenotypes: 9p minus syndrome, acromelic frontonasal dysostosis, amyotrophic lateral sclerosis, anophthalmia microphthalmia, attention-deficit/hyperactivity disorder, autism, bipolar disorder, Cantu syndrome, cerebral palsy, congenital diaphragmatic hernia, congenital heart disease, controls, developmental disorder, early onset Alzheimer’s disease, early onset Parkinson’s disease, epilepsy, intellectual disability, isolated biliary atresia, mixed, neural tube defects, neurodevelopmental disorder, nonsyndromic cleft lip / palate, obsessive compulsive disorder, orofacial cleft, preterm birth, periventricular nodular heterotopia, schizophrenia, sporadic congenital hydrocephalus, sporadic infantile spasm syndrome, syndromic craniosynostosis, Tourette syndrome, and vein of galen malformations. DNVs were extracted from each publication and, if necessary, lifted to build 38 of the human genome. All variants were annotated using Variant Effect Predictor ^32^. The VCF files, the annotated files, and a README file are on Zenodo (DOI 10.5281/zenodo.13901295).

To assess regions on the p-arm of chromosome 9 that have enrichment of DNVs, we retained only variants on the p-arm (b38 chr9:1-42668912). Since there are many variable phenotypes in 9p deletion and duplication syndromes ^1,2^, we retained DNVs from individuals with the following phenotypes for analyses in this study: acromelic frontonasal dysostosis, amyotrophic lateral sclerosis, anophthalmia microphthalmia, attention-deficit/hyperactivity disorder, autism, bipolar disorder, cerebral palsy, congenital diaphragmatic hernia, congenital heart disease, developmental disorder, early onset Alzheimer’s disease, early onset Parkinson’s disease, epilepsy, intellectual disability, isolated biliary atresia, neural tube defects, neurodevelopmental disorder, nonsyndromic cleft lip / palate, obsessive compulsive disorder, orofacial cleft, preterm birth, periventricular nodular heterotopia, schizophrenia, sporadic congenital hydrocephalus, sporadic infantile spasm syndrome, syndromic craniosynostosis, Tourette syndrome, and vein of galen malformations. There were a total number of 62,181 individuals within these phenotypes.

### DNVs from WGS from NDDs

We analyzed genomes from 5,824 individuals with autism and their parents from the Simons Simplex Collection ^33^ and SPARK ^34^. DNVs were called using HAT ^27^ and quality checked using acorn ^28^. Beyond standard DNV filtering in HAT, DNVs were additionally filtered and removed if they were in the following genomic regions: simple repeats and sdust that contains low complexity genomic regions ^35^.

### Genomic Regions for Testing DNV Enrichment

Different genomic annotations were used for testing enrichment of DNVs. These included 3’ UTRs, 5’ UTRs, promoters, and coding exons of RefSeq protein-coding genes. These annotations were downloaded from the UCSC Genome Browser ^36^ using the Table Browser tool. To assess putative, noncoding regulatory regions we examined candidate cis-regulatory elements (CREs) from the human fetal brain ^37^ using data from the Catlas database ^38^, CREs from all of ENCODE ^39^ (GRCh38-cCREs.bed) available at https://screen.wenglab.org/downloads, and the VISTA Enhancer Browser ^40,41^ CREs available at https://enhancer.lbl.gov/vista/downloads.

### Development of DiamondsDenovo

In this study, we developed a tool called DiamondsDenovo to assess the enrichment of DNVs within genomic regions. DiamondsDenovo is comprised of two subtools. The first subtool is a strategy to generate mutation rates for specified regions by comparing a reference genome from a species of interest to the human reference genome. Input data files to this program include the regions of interest in a bed file, the human reference genome, the other species reference genome, the divergence time between the two species, the chain file between the two reference genomes ^36^, and a bigwig ^36^ file of CADD scores ^42^. For each region, the tool identifies the region (if present) in the other species genome that corresponds to its location in the human genome. The sequence from each species is than extracted from both reference genomes. The sequences are then aligned using MUSCLE ^43^. From the sequence alignment, the aligned length, number of mutations, and mutation rate are calculated from the data. A weighted mutation rate is also calculated using the following formula: (mutation rate x (1/ average CADD score in the region)). A table containing this data is output by the program and is an input for the second part of DiamondsDenovo. The second subtool in DiamondsDenovo calculates a p-value for DNV enrichment in regions of interest. The p-value is calculated using the following formula in R: binom.test(x = DNV count, n = number of individuals * 70 DNVs per genome, p = mutation rate). It takes as input a bed file of DNVs, a bed file of regions of interest, a text file of mutation rates (calculated in subtool 1), CADD score bigwig, number of samples, and an option to use the CADD weights or not. The output gives the p-value for all the regions, a Manhattan plot, and a QQ plot. The Manhattan plot uses a genome-wide significant line of 8.33 × 10^−12^ (calculated as (0.05/9,000,000,000 nucleotides * 2)) to correct for all nucleotides in a diploid genome and a suggestive line of 5 × 10^−8^ (calculated as (0.05/1,000,000)). For conservative calculations in this study, we only use the unweighted mutation rates.

We chose two species for testing in this study and those include *Pan troglodytes* (chimpanzee) and *Takifugu rubripes* (fugu). Chimpanzee is a closely related species to human and has a divergence time of six million years. Fugu is a distant species with a divergence time of 450 million years, but it also has a condensed genome with the most critical genomic regions including in the noncoding genome ^44^. The names of each of the genomes used from the UCSC Genome Browser include hg38, panTro6, and fr3. For chimp-based and fugu-based testing of promoters, 3’ UTRs, 5’ UTRs, and coding exons we performed DiamondsDenovo using the 62,181 individuals from our updated denovo-db. For the CREs, we used the DNVs called from 5,824 individuals from the WGS datasets analyzed in this study.

### denovolyzeR

To test for enrichment of genes with an excess of DNVs in the 62,181 individuals from our updated denovo-db, we utilized the denovolyzeR tool ^45^ and tested the following categories: loss-of-function (lof), missense (mis), synonymous (syn), and lof+mis (prot). Genome-wide significance for a gene was set at 2.6 × 10^−6^.

### fitDNM

Another test to look for enrichment of DNVs within genomic regions was fitDNM ^46,47^. We ran fitDNM on the 62,181 individuals from our updated denovo-db for the promoters, 3’ UTRs, 5’ UTRs, and coding exons genomic regions. For the CREs, we used the DNVs called from 5,824 individuals from the WGS datasets analyzed in this study.

### Spatial Expression in E13.5 Mouse Head

A C57BL/6J wild type embryonic day 13.5 (E13.5) mouse head was collected from a wild type mouse at the Jackson Laboratory and processed through their *in vivo* services. Procedures were approved by the Institutional Animal Care and Use Committee of The Jackson Laboratory. Steps followed by the Jackson Laboratory services team included 1) the head was isolated and fixed in 4% PFA/PBS on ice for 50±15 min, 2) washed 3 × 5min in PBS on ice, 3) cryoprotected overnight in 30% sucrose/PBS at 2-8°C, 4) heads bisected median sagittally, and cryoembedded cut side down in OCT, and 5) cryoblocks stored at −80°C until shipment in dry ice. The tissue was then sent to MedGenome for sectioning and processing through the 10x Visium HD Spatial Gene Expression platform. Processing of the data was performed using a series of steps. First, the Space Ranger (version 2.0.1) mkfastq tool from 10x genomics was used to generate fastq files. Second, the Space Ranger count tool was used to analyze the microscope slide image and the fastq files. Specifically, the steps include alignment, tissue detection, fiducial detection, and counting of barcodes/UMIs. Third, the barcodes are used to generate feature-barcode matrices, identify clusters, and analyze gene expression. Several output files are generated including an HDF5 file containing the feature barcode matrix, QC images, downsampled images, and barcode locations. Fourth, Seurat (version 4.1.0) was utilized to perform QC and to further analyze gene expression. Cell QC was conducted using several metrics to ensure data integrity. The number of genes expressed (nFeature_Spatial) was assessed by counting genes with a UMI count greater than zero in each cell or spot, while the number of UMI reads (nCount_Spatial) represented the total UMI count from genes with at least one detected transcript per cell or spot. Mitochondrial contamination was evaluated by calculating the percentage of UMI counts originating from mitochondrial genes for each cell or spot within a sample. Similarly, ribosomal contamination was measured as the fraction of UMI counts attributed to ribosomal protein genes, which reflect translational activity rather than rRNA contamination. Since ribosomal gene abundance varies across cell types, this metric provides insight into data composition. Additionally, hemoglobin gene contamination was assessed to detect potential blood contamination. Fifth, once the data passed QC, the subsequent analyses were to normalize the data using SCTransform. A regularized negative binomial regression is used by this function to normalize the UMI count data. Sixth, the SCTranform function looks for features contributing to large cell-to-cell variation in the dataset. Seventh, the data is scaled using a linear transformation. Seventh, PCA is performed on the scaled data. Eighth, a K-nearest neighbor graph is the clustering approach used in Seurat to identify ‘communities’ using the first 14 principal components from the PCA. The FindClusters function optimizes to find the best clusters. Ninth, a UMAP is generated to explore the data. Tenth, the FindAllMarkers function is used to find gene markers specific to each cluster and it uses the Wilcoxon Rank Sum test.

### Assessment of Published Mouse Phenotypes

We utilized MouseMine ^48^ to query for known phenotypes, in mouse models, associated with twenty-four genes that we identified in this study as relevant to 9p deletion syndrome. Twenty-one genes had known phenotypes.

## RESULTS

In this study, we performed whole-genome sequencing on 100 individuals with 9p-related syndromes and family members. Most individuals sequenced were independent probands (n = 85), two were affected siblings, five were fathers, six were mothers, one was a grandfather, and one was a grandmother. This is the largest cohort of individuals ever assembled for 9p-related syndromes regarding whole-genome sequencing data and provides a unique resource to delineate core features of these syndromes.

### Genomic Architecture of 9p-Related Syndromes and Implications for Individuals

The genomic architecture of 9p genes in 9p-related syndromes was analyzed by examining their copy number, using an approach with paralog specificity, in all sequenced individuals in this study. Subsequently, data from the 1000 Genomes Project (controls) was processed in the exact same manner. The two datasets were combined, and we used principal component analysis to perform unsupervised learning on the data. The results are shown in Figure 1. In the center of the PCA are individuals from the 1000 Genomes Project who have copy numbers of two for most genes on 9p. On the left side of the plot and moving up the y-axis are individuals ordered by increasing deletion size starting from the telomere and getting larger as shown in orange. On the bottom left, is the one individual in our study who has a deletion near the centromere shown in orange. On the top right are three unrelated individuals who have a large deletion shown in orange followed by a large duplication shown in blue on the p-arm. Viewing the middle/bottom of the plot and moving to the right are individuals with increasing duplication size shown in blue starting with duplications that are approximately half the p-arm in size moving to those with full duplication of 9p. By using estimates of copy number with paralog-level specificity that is only derivable using whole-genome sequencing data the first complete view of the genomic architecture of 9p genes in 9p-related syndromes has been accomplished in this study. Future work and implementation of these results will enable more refined matching of individuals with 9p-related syndromes to other families. For example, the three unrelated individuals at the top right of the plot containing similar deletion/duplication events likely share more phenotypes with each other than they do with the individual at the bottom left who has a deletion near the centromere. The general clinical information for these four specific individuals says they have 9p deletion syndrome. However, whole-genome sequencing clearly shows these are not the same genomic events at all. Further genotype-phenotype analyses are required to fully determine the links of the events on this genomic map to specific phenotypes.

**Figure 1:**
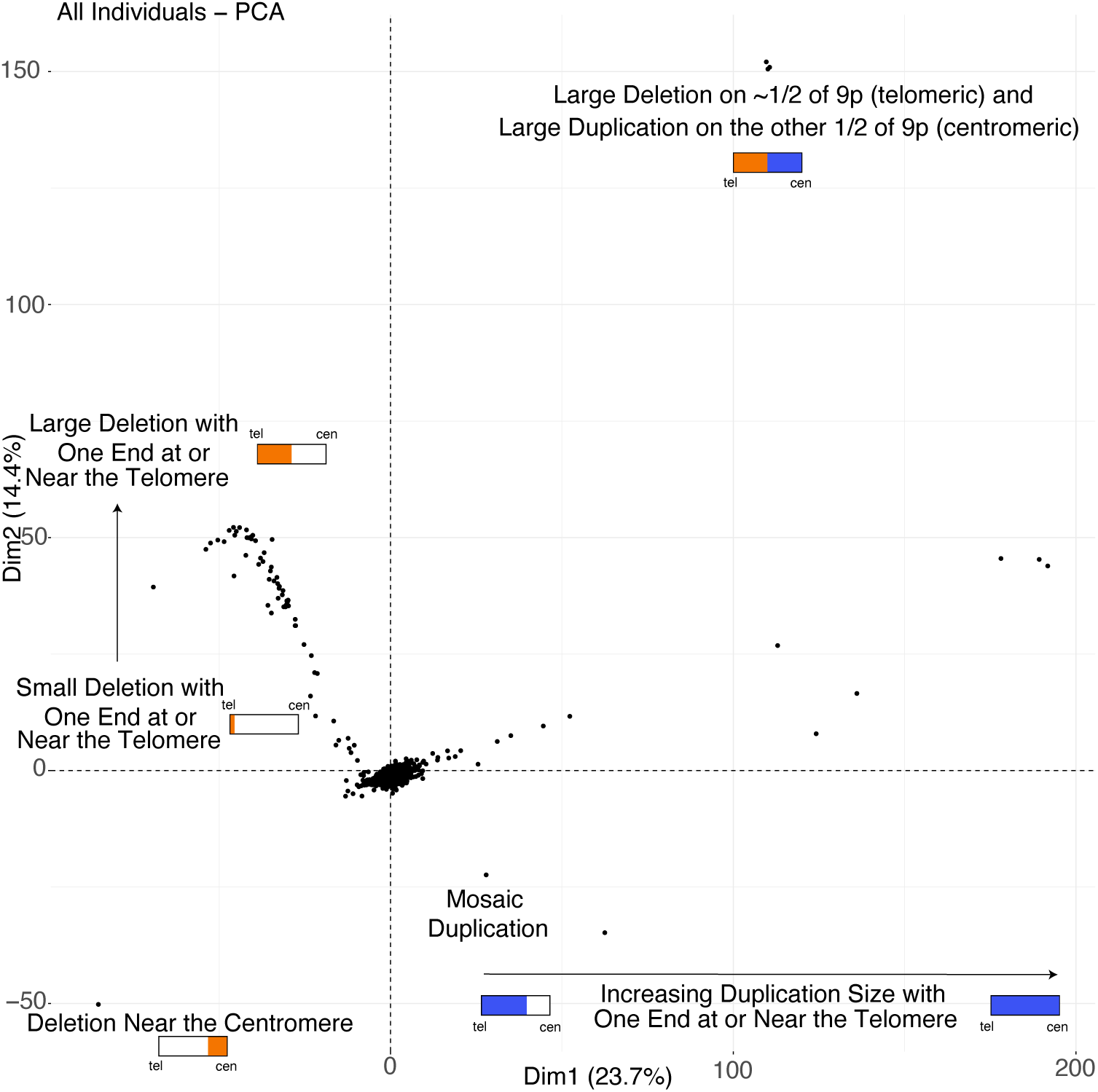
Genomic Architecture of 9p-Related Syndromes. Each point, in this principal component analysis of genic copy number on 9p, represents an individual either sequenced in this study as part of a family with 9p-related syndromes or from the 1000 Genomes Project (controls). The individuals from the 1000 Genomes Project cluster in the center of this plot. Small schematics of the p-arm are shown in rectangles with tel indicating telomere and cen indicating centromere. Orange represents deletion and blue represents duplication. This PCA reveals the complexity of what is called a 9p deletion syndrome (all the individuals with orange) and what is called a 9p duplication syndrome (all the individuals with blue). This kind of map is critical for researchers and families to connect with each other based on similar genomic events and for future refined care, beyond research, and into the clinic.

### Machine-Learning Model to Predict 9p Deletion Syndrome

Copy number of the 488 genes on 9p was calculated (as described above) in individuals with 9p deletion syndrome and in individuals from the 1000 Genomes Project (who do not have 9p deletion syndrome). A Random Forest machine learning approach was used to train a model using 70% of each dataset in the training set and 30% of each dataset in the testing set. Using 5-fold cross validation on the training set we found that our model had high accuracy of 0.997, Sensitivity of 0.9, Specificity of 1, Positive Predictive Value of 1, and a Negative Predictive Value of 0.997. The top 10 features (genes) contributing to the signal were *RFX3_DT* (Mean Decrease in Gini [MDG] = 5.67), *RCL1* (MDG = 4.81), *LOC105375956* (MDG = 4.57), *MIR101_2* (MDG = 3.30), *UHRF2* (MDG = 3.16), *SMARCA2* (MDG = 3.14), *RFX3* (MDG = 3.13), *RLN2* (MDG = 3.08), *LOC124902112* (MDG = 3.07), and *GLIS3* (MDG = 2.99). We also used a Naïve Bayes strategy to examine the 488 genes on 9p with training and testing set up in the same manner as in the Random Forest. The result of the Naïve Bayes strategy was an accuracy of 0.996, Sensitivity of 0.95, Specificity of 0.997, Positive Predictive Value of 0.904, and Negative Predictive Value of 0.999. The accuracy of this model indicates that we have good ability to identify individuals with 9p deletion syndrome using whole-genome sequencing data.

### Precision Genomics in 9p Deletion Syndrome

Most individuals sequenced in this study have 9p deletion syndrome. These individuals were assessed by a minimum of one sequencing technology and karyotype. Characterization of the structural variants in each individual revealed a total of 184 breakpoints. Precise nucleotide resolution was achieved in 159 (86.4%) of these events. Furthermore, we predicted ten translocation events from the sequencing data, and all were confirmed to be real by FISH. Eleven individuals contained structural variation involving chromosome 9 and at least one other chromosome. In one individual, the variation was predicted to be quite complex based on the karyotype and sequencing results (Figure 2) and is likely the result of chromothripsis^49^. Using breakpoint information from sequencing data, FISH probes were designed, and all predictions were confirmed by the FISH experiments. This emphasizes the importance of sequencing data and its accuracy in identification of fully resolved structural variation. Reports containing these schematics and other related information were conveyed back to the research participants. Overall, 53 individuals had deletion alone (75.7%), four had deletion and duplication on chromosome 9 (5.7%), 11 individuals had more complex variation also involving a translocation of sequence from another chromosome (15.7%), and two individuals had no large genic deletions (2.9%).

**Figure 2.**
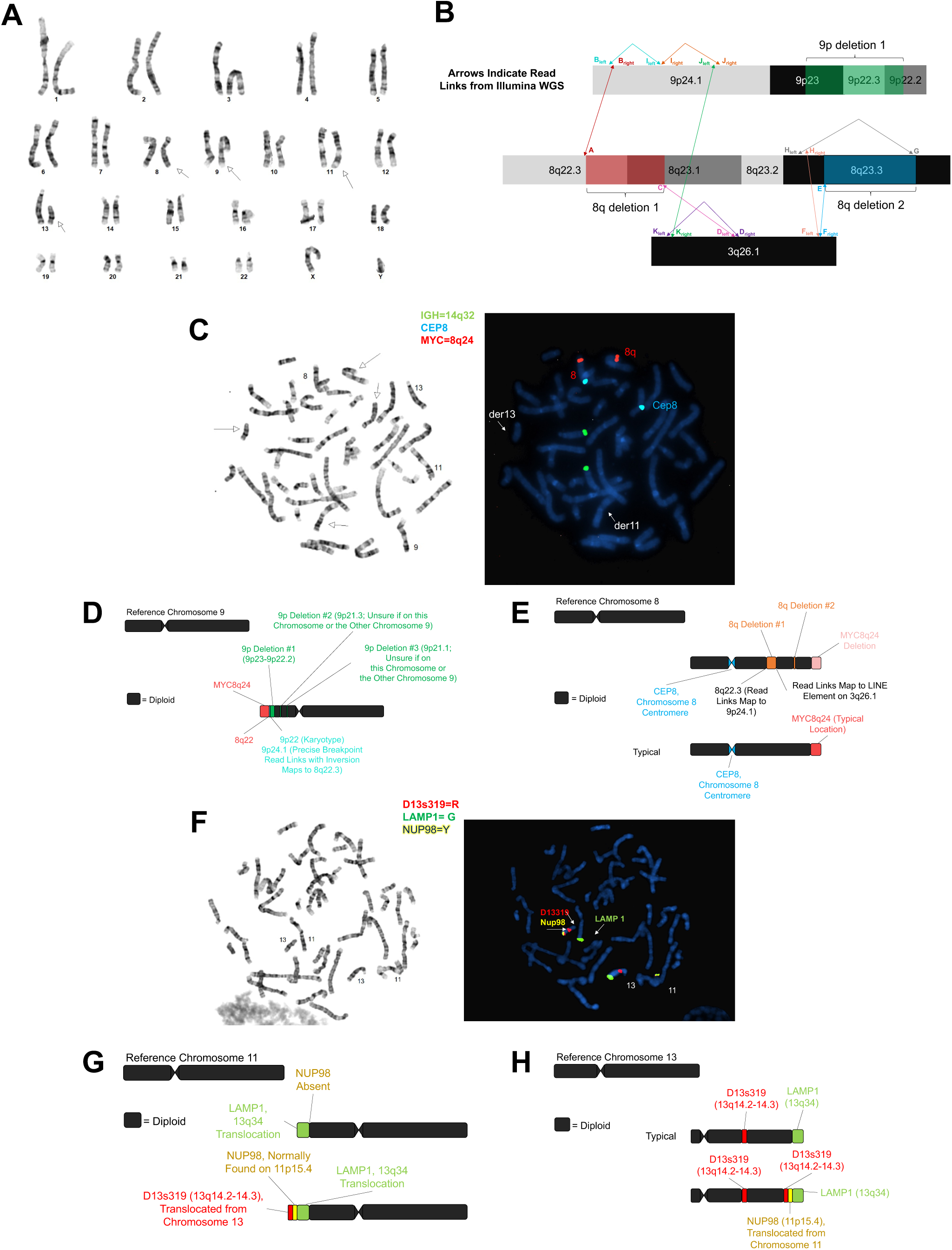
Complex Structural Variation Detected in 9p.123.p1. **A)** Shown is the karyotype of individual 9p.123.p1 where abnormalities were detected involving chromosome 8, chromosome 9, chromosome 11, and chromosome 13. **B)** Shown is the detailed characterization of the complex variation involving chromosome 8, chromosome 9, and chromosome 3 as detected using short-read whole-genome sequencing. A = 8q del 1 breakpoint 1 (chr8:105,028,001; linked to B_right_). B_right_ = right (centromere) side of region B (chr9:8,384,167; linked to A). B_left_ = left (telomere) side of region B; inverted reads at chr9:8,384,164; linked to I_left_. C = 8q del 2 breakpoint 2 (chr8:107,839,946; linked to D_left_). D_right_ = chr3:166,346,923 (linked to K_left_). D_left_ = chr3:166,346,922 (linked to C). E = 8q del 2 breakpoint 1 (chr8:114,357,399; linked to F_right_). F_right_ = right (telomere) side of region F (chr3:167,475,935; linked to E). F_left_ = left (centromere) side of region F (chr3:167,475,865; linked to H_right_). G = 8q del 2 breakpoint 2 (chr8:114,697,049; linked to H_left_). H_right_ = right (telomere) side of region H (chr8:111,368,737; linked to F_left_). H_left_ = left (centromere) side of region H (chr8:111,368,715; linked to G). I_right_ = inverted reads at chr9:8,430,297 (linked to J_right_). I_left_ = inverted reads at chr9:8,430,297 (linked to B_left_). J_right_ = inverted reads at chr9:8,719,693 (linked to I_right_). J_left_ = chr9:8,719,701 (linked to K_right_). K_right_ = chr3:157,869,331 (linked to J_left_). K_left_ = chr3:157,869,330 (linked to D_right_). Not to scale. **C)** Shown are the FISH results for chromosome 8 and chromosome 9. The green probe is to 14q32, the blue probe is to the centromere of chromosome 8, and the red probe is to 8q24. **D)** Shown is the updated schematic of chromosome 9 based on the karyotype, WGS, and FISH experiments. **E)** Shown is the updated schematic of chromosome 8 based on the karyotype, WGS, and FISH experiments. **F)** Shown are the FISH results for chromosome 11 and chromosome 13. The red probe is to 13q14, the green probe is to 13q34, and the yellow probe is to 11p15. **G)** Shown is the updated schematic of chromosome 11 based on the karyotype, WGS, and FISH experiments. **H)** Shown is the updated schematic of chromosome 13 based on the karyotype, WGS, and FISH experiments. Structural variation in this individual is likely due to chromothripsis.

We previously published DNVs detected in individual 9p.100.p1 using sequencing data generated in the father, mother, and child. In this study, we recruited three new trios and detected DNVs in these trios. In each case, the 9p structural variant event was *de novo*. Examining single-nucleotide variants and small insertions/deletions we did not find any missense or loss-of-function DNVs on 9p.

### Preferential Copy Number Breakpoints in Two Late-Replicating Regions of 9p

To understand the mechanism underlying the structural variants formed in 9p deletion syndromes, we examined published Repli-seq data ^50^ on the p-arm of chromosome 9 from ENCODE. Through this search, we identified four regions on 9p that are late-replicating regions (i.e., highest signal during S4 and G2) (Figure 3). We named these regions Late-Replicating Region 1 (LRR1, b38: chr9:7302516-14625487), Late-Replicating Region 2 (LRR2, b38: chr9:16012189-18667816), Late-Replicating Region 3 (LRR3, b38: chr9:22193299-26926755), and Late-Replicating Region 4 (LRR4, b38: chr9:27450126-32408022). These regions are shared across several cell types. Further review of these regions and comparison to the HumCFS fragile site database ^51^ identified LRR2 as the known human fragile site FRA9G that is on 9p22 and includes the gene *CNTLN* ^51,52^, LRR3 as the known human fragile site FRA9C and includes the gene *ELAVL2* ^51,53^, and LRR4 is the recently molecularly-mapped fragile site FRA9A with breakpoints spanning all 8.2 Mb of 9p21-1-9p21.3, caused by the (GGGGCC)N repeat expansion in C9orf72 ^51,54^. LRR1, including, *PTPRD*, resides in 9p24, a region associated with an un-named chromosomal fragile site ^55^ and a known recurrent DNA double-strand break cluster ^56^. Visual inspection revealed several breakpoints in LRR1 and LRR2 (Figure 3). There were no breakpoints in LRR3 or LRR4 identified in any individual with 9p deletion syndrome. We hypothesized that a replication-based mechanism may be critical for structural variant formation in 9p deletion syndrome. Of the 70 probands in our study, three had structural variants with a breakpoint in LRR1 and LRR2 (4.3%), 29 had structural variants with a breakpoint in LRR1 (41.4%), 11 had structural variants with a breakpoint in LRR2 (15.7%), 25 did not have breakpoints in LRR1 or LRR2 (35.7%), as mentioned above zero individuals had breakpoints in either LRR3 or LRR4, and two individuals had no 9p deletions (2.9%) (Figure 3). We further tested whether there was an enrichment of individuals with breakpoints at LRR1 or LRR2. For this, we calculated the proportion of 9p the regions encompass. LRR1 is 17.2% of the 9p region and LRR2 is 6.2%. In total, 32 individuals had a breakpoint in LRR1 and this was enriched given the size of the region (binomial test: n = 70, x = 32, p = 0.172, p-value = 3.0 × 10^−8^) and 14 individuals had a breakpoint in LRR2 that was also enriched given the size of the region (binomial test: n = 70, x = 11, p = 0.062, p-value = 3.8 × 10^−3^). These two regions are of critical importance in 9p deletion syndrome. One potential defined mechanism underlying the structural variants we identify in individuals with 9p deletion syndrome is chromothripsis^57–60^. This mechanism has been identified in some other congenital diseases ^61–64^.

**Figure 3.**
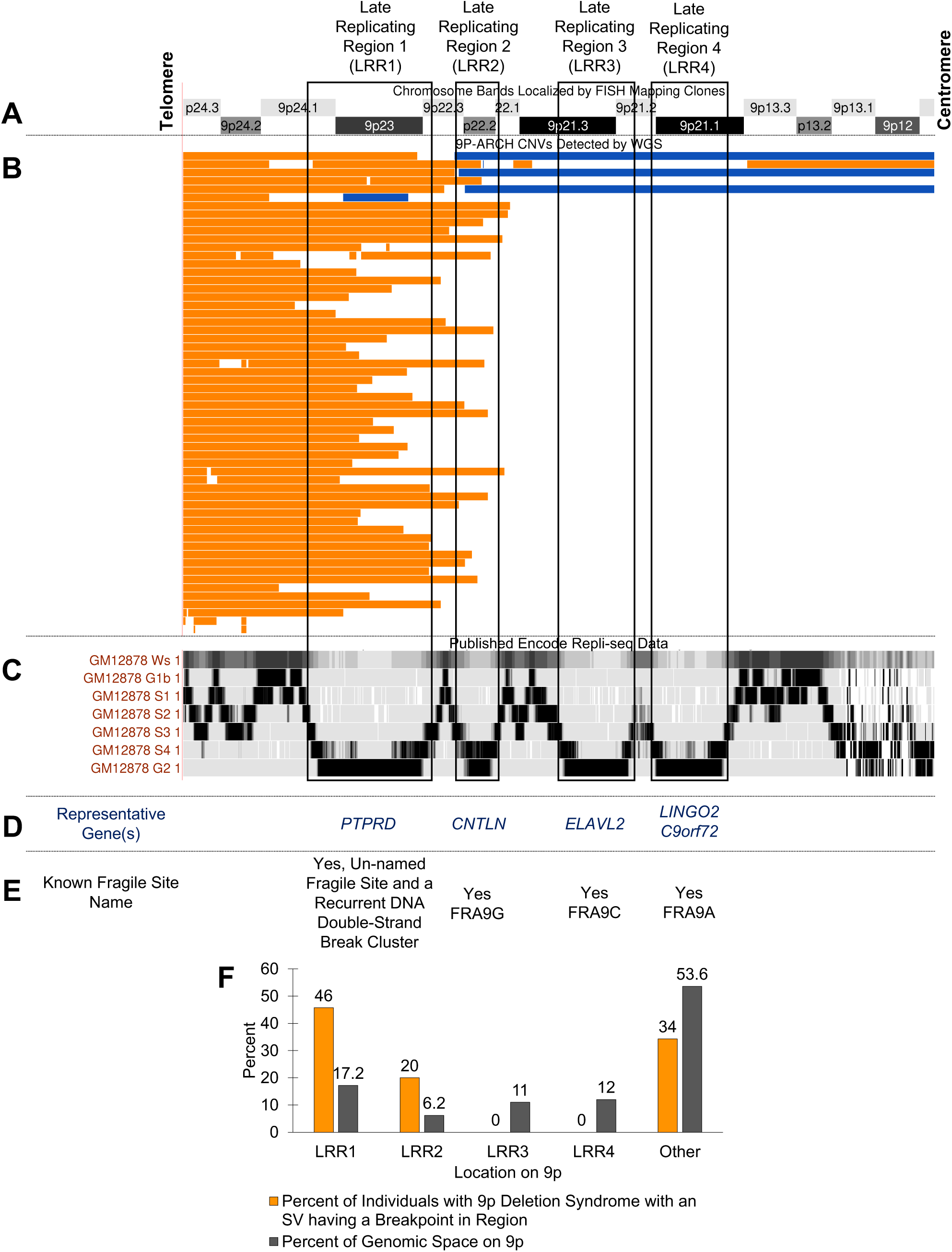
9P-ARCH Copy Number Variant Breakpoints Preferentially Localize to Two Late-Replicating Regions on 9p. A) Chromosome bands on the p-arm of chromosome 9. B) Shown in orange are deletions and in blue duplications identified by WGS in 9P-ARCH, respectively. C) The previously published Repli-seq tracks ^50^ are shown from ENCODE for GM12878 cells. These are consistent with replication timings in several other cell lines. We identified four late-replicating regions with highest signal intensity in the S4 phase or G2 phase of cell cycle. They are as follows: Late-Replicating Region 1 (LRR1, b38: chr9:7302516-14625487), Late-Replicating Region 2 (LRR2, b38: chr9:16012189-18667816), Late-Replicating Region 3 (LRR3, b38: chr9:22193299-26926755), and Late-Replicating Region 4 (LRR4, b38: chr9:27450126-32408022). D) and E) The majority of breakpoints identified in individuals with 9p deletion syndrome resided in LRR1 on 9p. This region contains the long gene (*PTPRD*) and is a region known to be an un-named fragile site and recurrent double-stranded break cluster region ^56^. Several of the other events have breakpoints in LRR2, which is the known human fragile site FRA9G. There are no breakpoints in the LRR3 or LRR4 that are known to be human fragile sites (FRA9C, FRA9A). The data in this figure support replication-based issues in LRR1 and LRR2 underlying the variation in 9p deletion syndromes. F) Shown is the percent of individuals with 9p deletion syndrome that have a breakpoint in one of the late replicating regions or in another part of 9p (orange) and the percent of genomic space that region takes up on 9p (gray). There is a significant excess of breakpoints in LRR1 (p = 3.0 × 10^−8^) and LRR2 (p = 3.8 × 10^−3^).

### Identification of Relevant 9p Genes in 9p Deletion Syndrome

Focused analyses of the copy number of genes on 9p was pursued in the individuals diagnosed with 9p deletion syndrome (n = 70) in comparison to control individuals from the 1000 Genomes Project (n = 2,504) who do not have a diagnosis of 9p deletion syndrome. For each gene on 9p, a Mann-Whitney U Test was performed to identify the genes with a significant difference in copy number between the two groups. Of the 488 genes, 287 (58.8%) showed a significant difference in copy number. By adding information on constraint from gnomAD ^14^ and probability of haploinsufficiency from a previous publication ^13^ (Figure 4), we found 60 of the 287 genes met some criteria for constraint or haploinsufficiency. It should be noted that 161 significant genes (most are LOC genes) on 9p do not have any constraint or dosage metrics available in the files we downloaded for constraint and probability of haploinsufficiency. For this reason, we examined additional evidence of potential relevance of significant 9p genes. Next, we looked for genes in this region with enrichment of DNVs on 9p, in two large cohorts, as another layer of information to prioritize genes.

**Figure 4:**
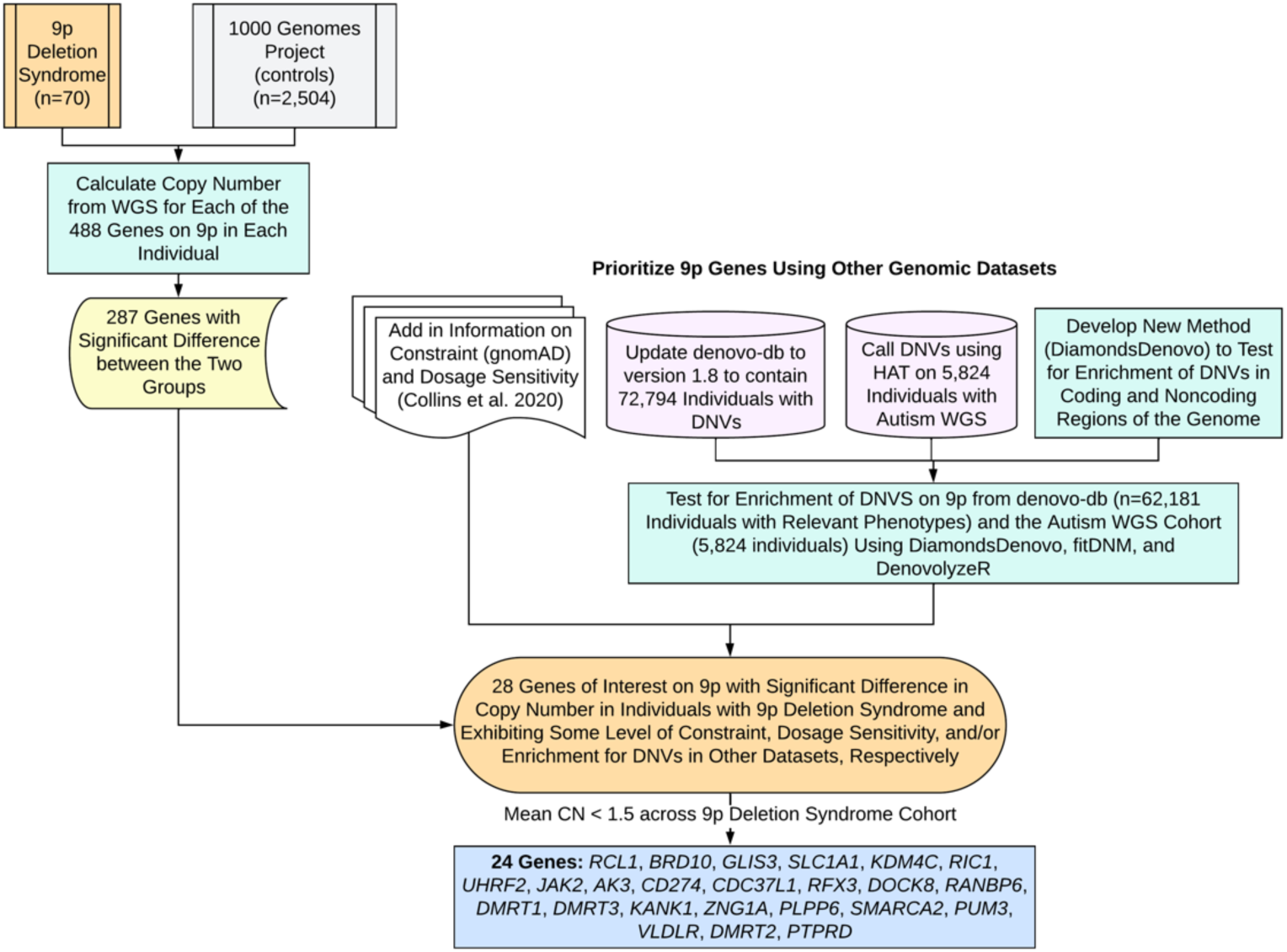
Prioritization of Genes in 9p Deletion Syndrome. Show is the gene prioritization approach in this study to reduce the number of total 9p genes from 488 down to 24 genes of interest. This approach required comparison of our sequencing data with control sequencing data. Integration of other genomic dataset including our expanded denovo-db and detection of DNVs in WGS data from families with autism was necessary. Next, we developed a new statistical method called DiamondsDenovo and used this in parallel to other tools (fitDNM, denovolyzeR) to find genes and genomic regions with excess DNVs in individuals with relevant phenotypes. Finally, the list was narrowed down further to 24 genes with mean copy number less than 1.5 across individuals with 9p deletion syndrome sequenced in this study.

Two cohorts were assessed for enrichment of DNVs on 9p (Figure 4). The first cohort was assembled through extensive searching of the literature, and we identified 40,198 unique DNV sites on the p-arm of chromosome 9 in 72,794 individuals. These DNVs are included in our newest release (version 1.8) of denovo-db. Testing for enrichment did not include individuals who did not have a phenotype of interest and the final dataset for testing consisted of 62,181 individuals. Since individuals underwent either whole-exome sequencing or whole-genome sequencing, we focused our enrichment of *de novo* variants in this dataset only on regions accessible by whole-exome sequencing. These included 5’ UTR, 3’ UTR, promoters, coding exons, or the entire gene (by category: synonymous, missense, LOF, missense+LOF). Within coding exons testing for enrichment of loss-of-function and missense DNVs was considered separately from testing for enrichment of synonymous DNVs. No genes had enrichment of DNVs in their 5’ UTR, one gene had enrichment in the 3’ UTR (*AK3*), no genes had enrichment in their promoter, 14 genes contained at least one coding exon with an excess of missense and LOF variants (*BRD10*, *CCIN*, *GLIS3*, *KANK1*, *KIF24*, *MOB3B*, *MYORG*, *RANBP6*, *RIC1*, *RUSC2*, *SMARCA2*, *TAF1L*, *TOPORS*, *UBAP1*), and no coding exons had an excess of synonymous variants. For the entire gene analyses, eight genes had an excess of synonymous DNVs (*ATOSB*, *BRD10*, *CIMIP2B*, *PUM3*, *RIC1*, *SAXO1*, *SPATA31F1*, *ZNG1A*), 16 genes had an excess of missense DNVs (*ATOSB*, *BAG1*, *BRD10*, *MYORG*, *PHF24*, *PLPP6*, *PUM3*, *RIC1*, *RIGI*, *SMARCA2*, *SPATA31F1*, *SPATA31F3*, *SPATA31G1*, *SPMIP6*, *WASHC1*, *ZNG1A*), seven genes had an excess of LOF DNVs (*BRD10*, *HACD4*, *MYORG*, *PHF24*, *PLPP6*, *RIC1*, *SAXO1*), and 19 genes had an excess of LOF+missense DNVs (*ATOSB*, *BAG1*, *BRD10*, *HACD4*, *MYORG*, *PHF24*, *PLPP6*, *PUM3*, *RFX3*, *RIC1*, *RIGI*, *SAXO1*, *SMARCA2*, *SPATA31F1*, *SPATA31F3*, *SPATA31G1*, *SPMIP6*, *WASHC1*, *ZNG1A*). The second cohort of individuals assessed were 5,824 individuals with autism with available whole-genome sequencing data from SPARK or SSC. We detected 4,066 DNVs on the p-arm of chromosome 9 in this data. We focused our analyses of this data to regions of the noncoding genome including candidate cis-regulatory elements in fetal brain, generally in all ENCODE, and in VISTA enhancers. We did not identify regions meeting genome-wide significance in this data. It should be noted that 5,824 individuals are substantially fewer than the 62,181 individuals available for the coding region analyses. Future increase in sample size would help with detection of relevant noncoding regions. For example, the candidate cis-regulatory element with the lowest p-value in the data (b38: chr9:12904581-12904931) contained three different DNVs and had a p value of 8.80 × 10^−10^ but did not reach our conservative cutoff of p < 8.33 × 10^−12^.

By adding information regarding significant genes from the *de novo* variant analyses, we expanded our list of 9p genes, with potential relevance, to 71 genes. Since many deletion cases are within the range of the telomere to 9p22 cytobands, we focused in specifically on the subset of these genes within these chromosome bands. Through these analyses, the set of genes for consideration was 28 (*RCL1*, *BRD10*, *GLIS3*, *SLC1A1*, *KDM4C*, *RIC1*, *UHRF2*, *JAK2*, *AK3*, *CD274*, *CDC37L1*, *RFX3*, *DOCK8*, *RANBP6*, *DENND4C*, *DMRT1*, *PSIP1*, *DMRT3*, *KANK1*, *ZNG1A*, *PLPP6*, *SMARCA2*, *PUM3*, *VLDLR*, *DMRT2*, SLC*24A2*, *PTPRD*, *BNC2*). Twenty-four of these genes exhibited an average copy number less than 1.5 for the 9p deletion cohort (*RCL1*, *BRD10*, *GLIS3*, *SLC1A1*, *KDM4C*, *RIC1*, *UHRF2*, *JAK2*, *AK3*, *CD274*, *CDC37L1*, *RFX3*, *DOCK8*, *RANBP6*, *DMRT1*, *DMRT3*, *KANK1*, *ZNG1A*, *PLPP6*, *SMARCA2*, *PUM3*, *VLDLR*, *DMRT2*, *PTPRD*). Based on genomic analyses alone, this list of genes is the best candidate gene list attainable at this time for 9p deletion syndrome as they are deleted in up to 83% of individuals with this syndrome.

### Identification of Expression Patterns of 9p Genes

Since neurodevelopmental disorders are a shared feature across 9p deletion syndrome, a spatial transcriptomic experiment of an embryonic mouse head at E13.5 was performed as this is a critical time in embryonic brain development including expansion of the forebrain and refinement of the midbrain and hindbrain. By examining the whole head, we also were able to gain information regarding craniofacial development as this is a time when the upper jaw, nose, and lower jaw are being formed. In Figure 5, we show the expression patterns in the mouse head with several genes showing expression in both the face and the brain. Both regions are relevant in relation to 9p-related syndromes. While several genes exhibit expression in the face, two were in very specific regions of the face and those included *Glis3* and *Bnc2*. Biallelic loss of function of *GLIS3* is known to underlie a syndrome in humans with several facial dysmorphic features ^65^. In the brain, some genes have restricted expression to one area (e.g., *Dmrt3* in radial glial cells) whereas there are several genes with expression in numerous regions of the brain (*Uhrf2*, *Cdc37l1*, *Rfx3*, *Psip1*, *Ptprd*, *Ak3*, *Smarca2*, *Pum3*).

**Figure 5:**
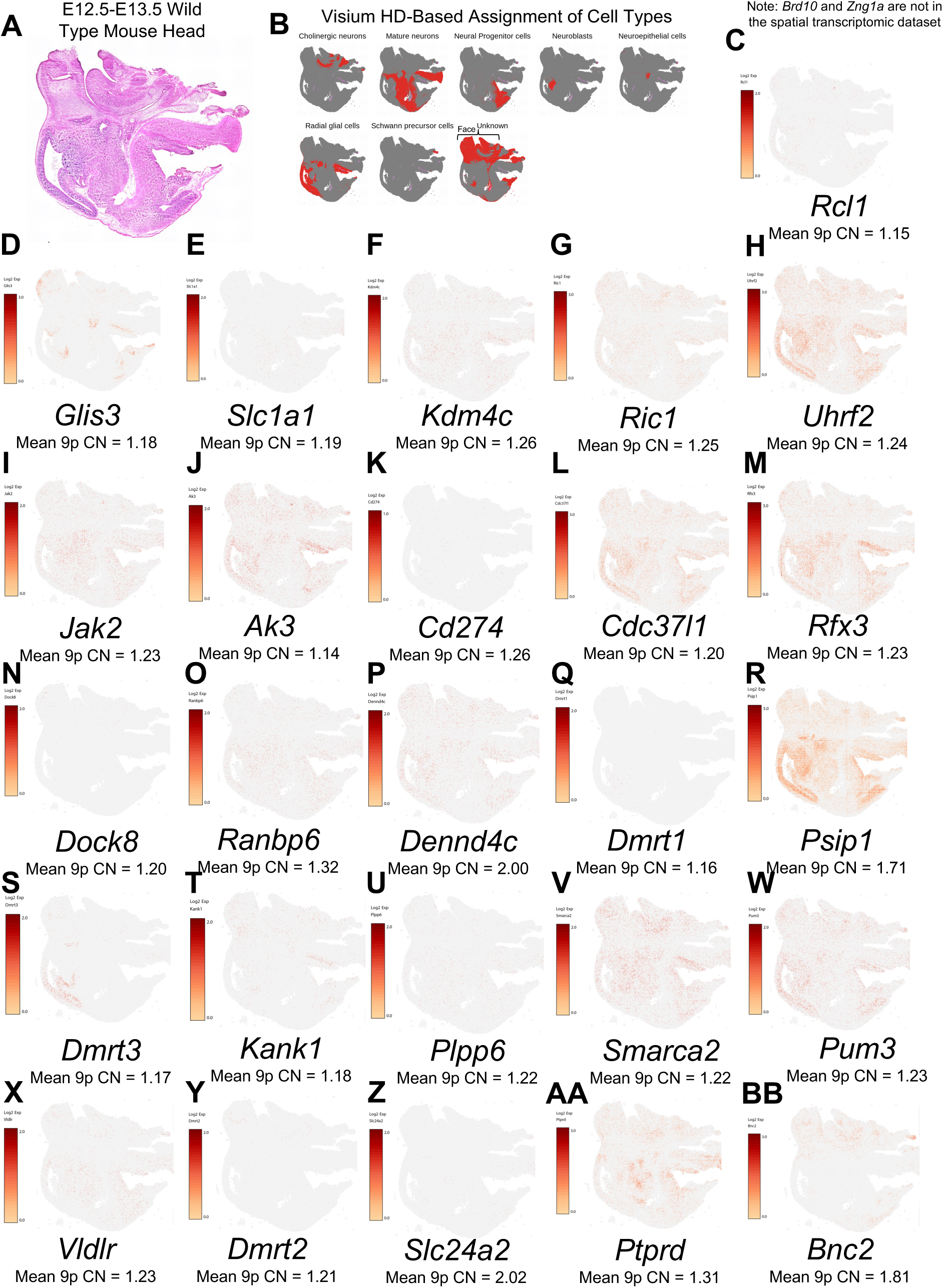
Spatial Expression in E13.5 Mouse Head of Genes Relevant to 9p Deletion Syndrome. A) Histology showing sagittal section of E13.5 mouse head. B) Cell type assignment of cells in 10X Visium HD experiment generated in this study. C) through BB) show the expression of the gene listed below each image. To the left of each image is the scale bar and below the gene name is the mean copy number (CN) of the gene in individuals with 9p deletion syndrome. Note, we originally looked to assess the full set of 28 genes but two were not represented in the Visium HD probe set (*Brd10*, *Zng1a*). The most interesting genes are the 24 genes where the mean 9p CN is < 1.5 across the 9p deletion syndrome cohort.

### Excess Mitochondrial Genome Copy Number in 9p

Two of the genes we identified as relevant in 9p deletion syndrome have consequences on mitochondrial function (*AK3*, *ZNG1A*). Therefore, assessment of the mitochondrial genome was pursued. For each individual, the mitochondrial genome was derived from whole-genome sequencing data. The mitochondrial genomes displayed expected relationships between mothers and their children (Figure 6). Testing of mitochondrial genome copy number was also pursued in both the individuals with 9p deletion syndrome in this study (n = 69 individuals) and individuals who are not affected with 9p deletion syndrome from a previous study ^66^ (n = 1,801 individuals). Only individuals with whole-genome sequencing data generated on DNA derived from *blood* were considered in this analysis. Individuals with 9p deletion syndrome had mitochondrial genome copy number of 271.4 ± 58.7 and individuals without 9p deletion syndrome had mitochondrial genome copy number of 219.5 ± 32.4. This was a significant difference in copy number (Mann Whitney U test p = 3.9 × 10^−15^) (Figure 6).

**Figure 6:**
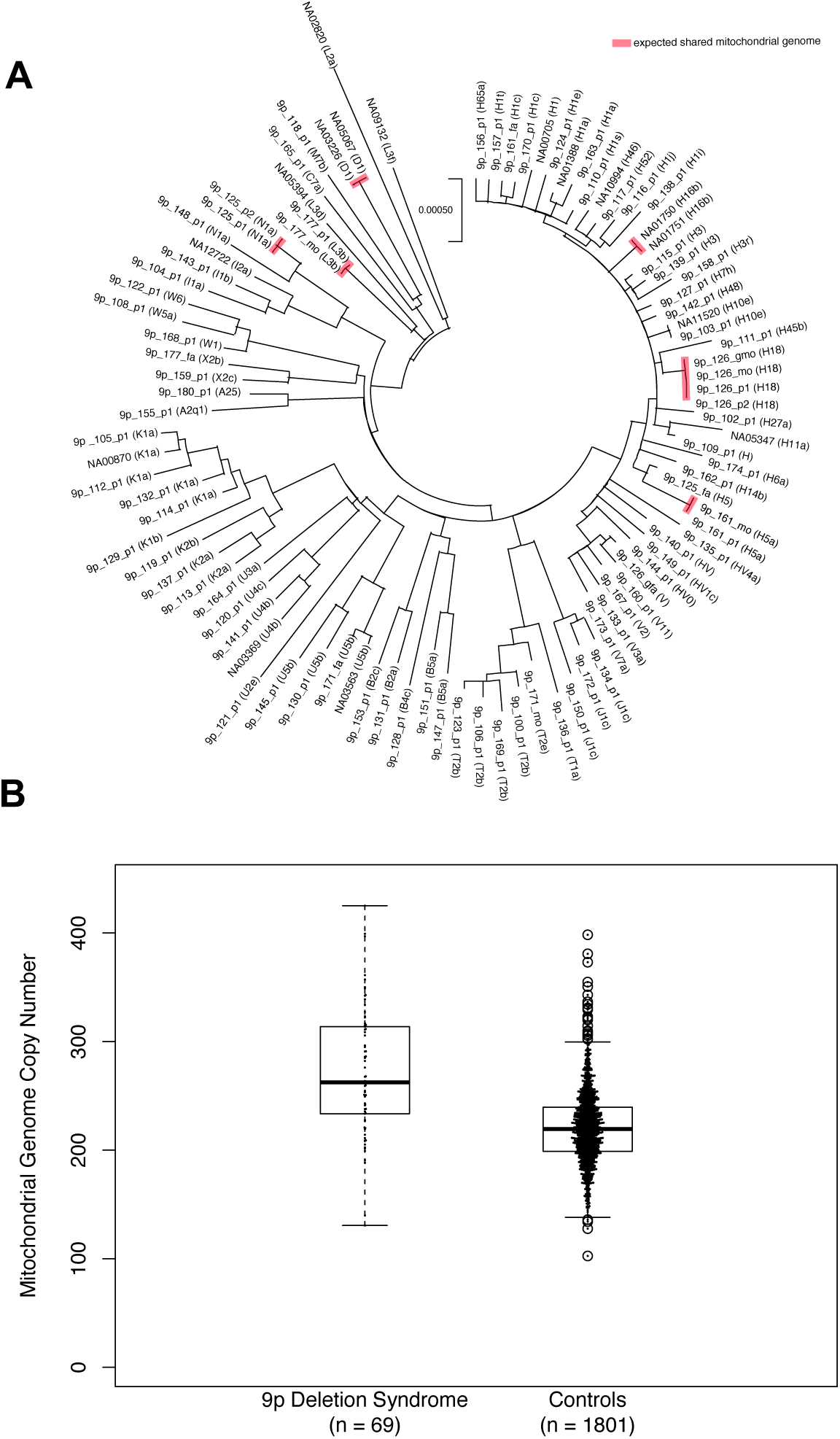
Mitochondrial Genome Assessment Using EGP on Illumina Short-Read Whole-Genome Sequencing Data. A) Maximum likelihood tree of mitochondrial genomes for all individuals sequenced in this study. The evolutionary history was inferred by using the Maximum Likelihood method and Tamura-Nei model. The tree with the highest log likelihood (−29132.73) is shown. Initial tree(s) for the heuristic search were obtained automatically by applying Neighbor-Join and BioNJ algorithms to a matrix of pairwise distances estimated using the Tamura-Nei model and then selecting the topology with superior log likelihood value. The tree is drawn to scale, with branch lengths measured in the number of substitutions per site. This analysis involved 98 nucleotide sequences. Codon positions included were 1st+2nd+3rd+Noncoding. There was a total of 16569 positions in the final dataset. Analyses were conducted in MEGA X. All familial relationships are confirmed in this analysis and individuals NA05067 and NA03226 are identified as the same individual. B) Mitochondrial genome copy number estimates specifically for individuals with WGS on *blood*-derived DNA. The data for the 1,801 controls is from 1,801 unaffected children that had previously been analyzed with EGP ^66^. There is a significantly higher mitochondrial genome copy number in individuals with 9p deletion syndrome (271.4 ± 58.7) than in controls (220.7 ± 32.4) (Mann Whitney U p = 3.9 × 10^-15^).

## DISCUSSION

The field of genomics is shifting towards whole-genome sequencing as a first-line approach to assess human phenotypes. However, syndromes due to large structural variants are often still assessed using lower resolution strategies including karyotypes and chromosome microarrays. Whole-genome sequencing provides fine scale resolution of the genome and is a needed next step as a standard for characterizing these syndromes. This study provides a first-of-its-kind look at 9p-related syndromes by performing whole-genome sequencing on 100 individuals in families with 9p-related syndromes, from both the WashU cohort collection and Coriell Repository, including 85 total unrelated probands with 9p-related syndromes to generate the clearest evaluation of variation in these syndromes. In Figure 1, a map of these syndromes shows the diversity at the level of the variant types in different individuals providing a glimpse into the potential reason for high phenotypic variability in these syndromes. This map will be used to link families to nearest other families in 9p gene space. Currently, strategies like GeneMatcher ^67^ enable researchers to link based on shared small variants in single genes. 9p-related syndromes now can have their own type of “CNVmatcher” within the syndrome’s genomic space. Furthermore, we generated a machine learning model to classify 9p deletion syndrome with high accuracy.

Whole-genome sequencing has enabled the resolution of complex structural variation in 9p-related syndromes including the complex case shown in Figure 2, which is likely due to chromothripsis. Gene level resolution is not attainable without whole-genome sequencing. We advocate for whole-genome sequencing as a first test for the assessment of these syndromes with addition of other orthogonal technologies (e.g., FISH, karyotype) as needed for confirmation analyses.

For future work at the level of the individual, 26 additional individuals with 9p-related syndromes were found with array data from various clinics. Future studies will include whole-genome sequencing of these individuals. Another future step is to sequence full trios especially for examining balanced translocation carrier parents with requisite consequences for genetic counseling in those families. In total, we have sequenced four complete trios including a child with a 9p deletion. One of these was previously published ^27^ and three were new to this study. Calling of DNVs was performed in this study and while no single-nucleotide or small insertion/deletion DNVs of obvious functional consequence on chromosome 9 were identified in these individuals, this set is a first-of-its-kind trio-based analyses of DNVs in whole-genome sequencing data in 9p-related syndromes. This is only the first step of what can be achieved using family-based WGS in these syndromes.

Cross-cohort analyses of 9p-related syndromes in relation to gene discovery was focused specifically on individuals with 9p deletion syndrome since most individuals sequenced in our study had this specific syndrome. Several strategies were used to prioritize the relevant genes for this syndrome (Figure 4) including major expansion of our database of DNVs and development of a new statistical method (i.e., DiamondsDenovo) to test for DNV enrichment on 9p in related phenotypes. In total, we identified 28 genes of interest and further characterized these by spatial expression experiments examining their expression in the developing mouse face and brain (Figure 5). Twenty-four of the genes (Figure 7) have an average copy number less than 1.5 across all individuals with 9p deletion syndrome and we next describe each of them further. *RCL1* is RNA terminal phosphate cyclase like 1. Copy number variants in this gene have been detected in multiple individuals with neurodevelopmental or psychiatric disorders ^68,69^. Some aspects of its expression pattern in the brain are unique to higher-order primates ^69^. It forms a complex with BMS1 to process 18s rRNA ^70^. *BRD10* is bromodomain containing 10 and is a gene that was formerly known as *KIAA2026*. Little is known about this specific protein, but it is likely based on its predicted function that it works as a regulator of transcription. *GLIS3* is GLIS family zinc finger 3. This gene has been identified as causal in autosomal recessive neonatal syndrome of diabetes mellitus and congenital hypothyroidism. It localizes to the nucleus and the primary cilia of cells ^71^. Expression of this gene was identified in the face and the brain in the present study. Individuals with the human syndrome have shared common facial dysmorphic features, including bilateral low-set ears, a depressed nasal bridge with an overhanging columella, elongated and upslanted palpebral fissures, a long philtrum, and a thin vermilion border of the upper lip ^65^. Many of these features are also observed in individuals with 9p deletion syndrome ^1,2^. However, the syndrome associated with this gene is autosomal recessive so more detailed genotype-phenotype studies are required to explore this further. *SLC1A1* is solute carrier family 1. In a five-generation family, the genetic variant responsible for schizophrenia and bipolar schizoaffective disorder was identified as a deletion specifically in this gene ^72^. Copy number variants in this gene have also been associated with schizophrenia ^73^. This gene also underlies autosomal recessive dicarboxylic aminoaciduria ^74^. It has also been associated with obsessive compulsive disorder ^75^. Specific missense variants in the gene underlie hot water epilepsy ^76^. It has also been suggested as a gene for autism based on linkage to 9p24 ^77^. Mice completely lacking this gene have been found to have age-dependent changes in behavior and in the brain. The effects include neurodegeneration and neuronal oxidative stress and has been suggested to be involved in Parkinson’s disease ^78,79^. *KDM4C* is lysine demethylase 4C (previously known as *JMJD2C),* which was associated with autism, obsessive compulsive disorder ^80^ and cancer ^81^. *RIC1* is RIC1 homolog, RAB6A GEF complex partner 1. It underlies the autosomal recessive condition called CATIFA syndrome ^82^. This syndrome is a rare neurodevelopmental disorder that is typified by distinctive facial features including long philtrum, elongated face, and small ears. There are also often vision issues and oral malformations including cleft lip and/or palate ^82,83^. *UHRF2* is ubiquitin like with PHD and ring finger domains 2. The gene is known for its role in cancer ^84^. In our spatial transcriptomic analyses, we found expression of this gene in many regions of the brain, including the radial glial cells, and in the face. A recent study of mice lacking this gene found several consequences on brain function and learning and memory ^85^. Another study of knockout mice showed the develop frequent seizures in adulthood with accompanying electrical activity abnormalities. They also found alterations in 5-methylcytosine levels in the brains of these mice ^86,87^. *JAK2* is Janus kinase 2. It is involved in several cancers and hematopoietic disorders ^88^. *AK3* is adenylate kinase 3. It has been found to localize to the mitochondrial matrix within cells ^89^. AK3 phosphorylates adenosine monophosphate using guanosine-5’-triphosphate (GTP) and is critical for the citric acid cycle ^90^. *CD274* is CD274 molecule. It is a gene implicated in cancer ^91^ and in the immune system and inflammatory response ^92^. *CDC37L1* is cell division cycle 37 like 1, Hsp90 cochaperone. It has expression throughout the brain in our spatial transcriptomics experiment. It has been indicated as a gene involved in cancer ^93^ and in regulating tau as part of tauopathies ^94^, respectively. *RFX3* is regulatory factor X3. In our spatial transcriptomics experiment, it is expressed in many places in the brain. It is well-established as a gene involved in autism ^95^ and is a regulator of cilia ^96–99^. *DOCK8* is dedicator of cytokinesis 8. It has been found as the gene involved in the autosomal recessive Hyper-IgE syndrome with recurrent infections ^100^ for which hematopoietic stem cell transplantation has been recommended as a potential therapy ^101^. It has also been identified as a regulator of microglia and potentially involved in neurodegeneration ^102^. *RANBP6* is RAN binding protein 6 and little is known about this gene. *DMRT1, DMRT2,* and *DMRT3* are part of a gene family and are called doublesex and mab-3 related transcription factor 1, 2, and 3, respectively. They are genes involved in sexual development ^103^. In our spatial transcriptomic analyses, *Dmrt1* and *Dmrt3* were expressed in the radial glial cells. The DMRT gene family has been indicated in brain development ^104^. *KANK1* is KN motif and ankyrin repeat domains 1. Variants in this gene have been identified in individuals with cerebral palsy ^105^, autism ^106^ nephrotic syndrome ^107^, or male congenital genitourinary anomalies ^108^. *ZNG1A* is Zn regulated GTPase metalloprotein activator 1A. It is a gene involved in the function of mitochondria and functions in the utilization of Zinc. In mice lacking *Zng1*, there is an impairment of the homeostasis of zinc and effects of Zinc in the diet. There is also an effect on mitochondrial function ^109^. *PLPP6* is phospholipid phosphatase 6. Little is known about this gene beyond that knockout mice exhibit effects related to allergies and lung function ^110^. *SMARCA2* is SWI/SNF related BAF chromatin remodeling complex subunit ATPase 2 and mutations result in autosomal dominant conditions Blepharophimosis-impaired intellectual development syndrome ^111^ and Nicolaides-Baraitser syndrome ^112^. Since this gene underlies a dominant disorder and some features overlap with 9p deletion syndrome it is likely a contributor to this syndrome. *PUM3* is pumilio RNA binding family member 3 and little is known about its function. *VLDLR* is very low-density lipoprotein receptor. It is the gene involved in autosomal recessive cerebellar hypoplasia impaired intellectual development, and dysequilibrium syndrome 1 ^113^. It is a receptor of reelin and important for brain development ^114^. Early work on *VLDLR* highlighted its role in lipid metabolism ^115^. *PTPRD* is protein tyrosine phosphatase receptor type D with specific expression in the brain ^116^. Mice completely lacking *Ptprd* (homozygous null) have learning problems ^116^. Furthermore, this gene is the orexigenic receptor for the asprosin hormone and mice completely lacking the gene are lean exhibiting appetite loss because they cannot respond to asprosin ^117^. Many of these genes have relevant phenotypes regarding 9p-related syndromes. Since two of them have links to mitochondrial dysfunction, testing of the mitochondrial genome was performed and individuals with 9p deletion syndrome have excess mitochondrial genome copy number in comparison with unaffected individuals.

**Figure 7.**
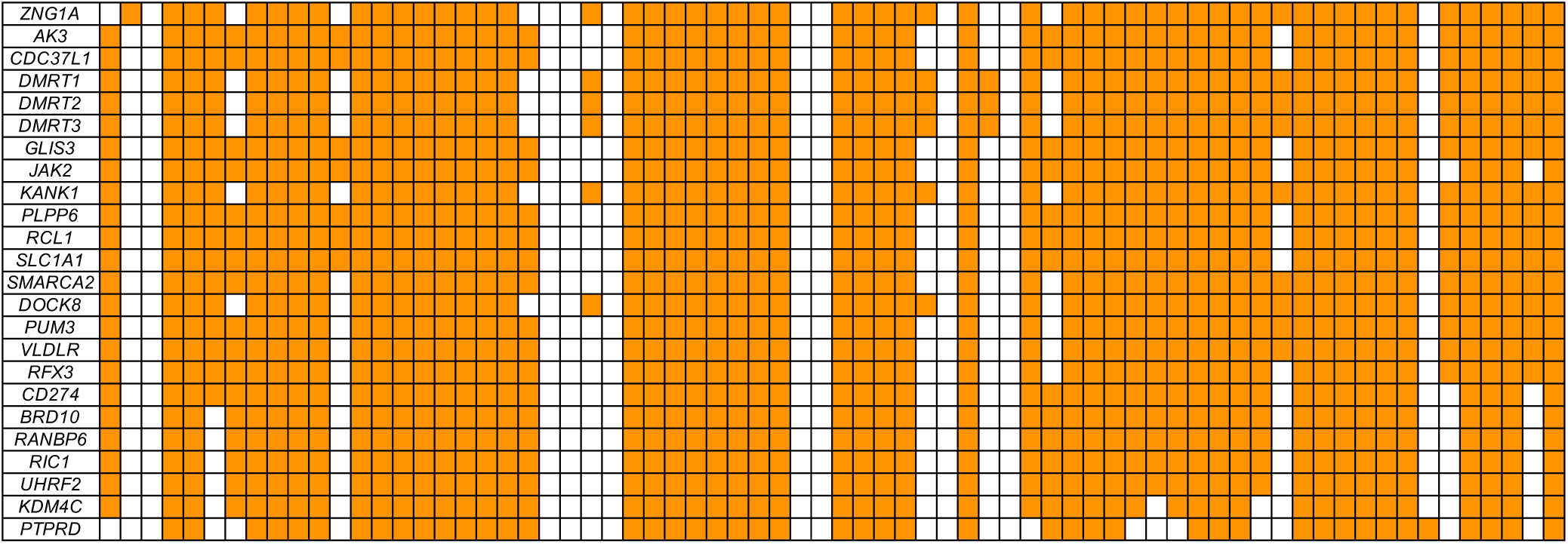
Copy Number Status of 24 Top Priority Genes in 9p Deletion Syndrome. Each box represents the copy number across the entire gene (rows) for a given individual (columns). Orange means the individual has a deletion that gene and white means they do not. The genes are sorted by the most individuals containing a deletion in the gene to the least. Note that smaller deletions within these genes are not represented in these estimates.

Whole-genome sequencing is an important tool to understand the genomics of human phenotypes. While it has not traditionally been a first line test for genomic syndromes including 9p-related syndromes, we show through this research study its utility. First, we built a comprehensive map of the genomic architecture of these syndromes using highly accurate gene copy number estimates from sophisticated analyses of whole-genome sequencing data (Figure 1). Our first use of this data is to relay to families (who are interested), other families with similar events based on these estimates. We hypothesize that individuals close in the map will share more phenotypes than those who are distant from each other. Future studies will integrate phenotypic data with information from this genomic map. As we show, 9p-related syndromes can be quite heterogenous with a minority of individuals having quite different genomic events than the majority. A CNVMatcher (akin to GeneMatcher) would be useful for researchers. Next, we built a machine learning model to classify 9p deletion syndromes using copy number estimates for genes on 9p. These models work well on our data and future studies will expand on this work. We highlight a particularly challenging case from the genomic perspective in Figure 2. While confirmations were performed using FISH, the predictions made by analyzing the sequencing data were all correct and resolved the variation in this individual. The second half of the paper focused on what critical genes underlie 9p deletion syndromes. We focused on individuals with a diagnosis of 9p deletion syndrome because the majority of our probands were from this group (n=70). By employing several techniques, we centered on 24 genes of critical importance and underlying 83% of individuals with 9p deletion syndrome. Some are well known and make sense regarding 9p-related phenotypes (e.g., *RFX3*, *SMARCA2*), while others are relatively enigmatic as to their function and phenotypic consequences of their disruption (*BRD10*, *GLIS3*, *RANBP6*, *PLPP6*, *PUM3*). Future work should address these 24 genes to further bring clarity to these syndromes.

## Data Availability

Code for DiamondsDenovo is available at https://github.com/TNTurnerLab/DiamondsDenovo. Sequencing data is currently under submission at AnVIL. Denovo-db version 1.8 is available at DOI 10.5281/zenodo.13901295.

https://doi.org/10.5281/zenodo.13901296

## ACKNOWLEDGMENTS

Thank you to the Chromosome 9P Minus Network and all the research participants who participated in this study. This work was supported by grants from the National Institutes of Health (R00MH117165 to T.N.T., R01MH126933 to T.N.T., R03HD116062 to T.N.T., P50HD103525 to T.N.T.), the Washington University in St. Louis Just-In-Time Core Usage Funding Program (JIT896 to T.N.T., JIT1174 to T.N.T.), the Simons Foundation (734069 to T.N.T.), Washington University in St. Louis laboratory startup funds to T.N.T., and a gift from Leon Eidelman and Sara Israel to the McDonnell Genome Institute. Thank you to Dr. Nara Sobreira, Dr. Joon-Yong An, Dr. Hee Jeong Yoo, Dr. Maria Chahrour, and Dr. Bryn Dionna Webb for searching your databases for additional individuals with 9p-related syndromes. The following cell lines/DNA samples were obtained from the NIGMS Human Genetic Cell Repository at the Coriell Institute for Medical Research: GM00705, GM00870, GM01388, GM01750, GM01751, GM02820, GM03226, GM03369, GM03563, GM05067, GM05347, GM05394, GM09132, GM10994, GM11520, GM12722. Thank you to Eunice Horton at the Coriell Institute for Medical Research for help with obtaining the DNA for these Coriell samples. Thank you to the New York Genome Center for generating the SSC and SPARK whole-genome sequencing data through the CCDG National Institutes of Health grant (UM1HG008901). We are grateful to all of the families at the participating Simons Simplex Collection (SSC) sites, as well as the principal investigators (A. Beaudet, R. Bernier, J. Constantino, E. Cook, E. Fombonne, D. Geschwind, R. Goin-Kochel, E. Hanson, D. Grice, A. Klin, D. Ledbetter, C. Lord, C. Martin, D. Martin, R. Maxim, J. Miles, O. Ousley, K. Pelphrey, B. Peterson, J. Piggot, C. Saulnier, M. State, W. Stone, J. Sutcliffe, C. Walsh, Z. Warren, E. Wijsman). We appreciate obtaining access to SSC whole-genome sequencing data on SFARI Base. Approved researchers can obtain the SSC population dataset described in this study by applying at https://base.sfari.org. We are grateful to all the families in SPARK, the SPARK clinical sites and SPARK staff. We appreciate obtaining access to SPARK whole-genome sequencing data on SFARI Base. Approved researchers can obtain the SPARK population dataset described in this study by applying at https://base.sfari.org. Thank you to Dr. Tristan Hayeck and Dr. Andrew Allen for previously developing and publishing fitDNM. Thank you to The Jackson Laboratory Model Generation Services and Breeding Services for creating and breeding the mice, and The Jackson Laboratory Preclinical Services for harvesting and processing the mouse samples.

## AUTHOR CONTRIBUTIONS

**Y.W.**: Formal analysis, Investigation, Writing – Review & Editing, Visualization. **E.I.S.**: Formal analysis, Investigation, Writing – Review & Editing, Visualization. **R.S.**: Formal analysis, Investigation, Writing – Review & Editing, Visualization. **S.C.**: Formal analysis, Investigation, Writing – Review & Editing, Visualization. **E.C.H.**: Formal analysis. **S.T.**: Investigation. **Y.C.H.**: Investigation. **C.M.**: Investigation. **K.V.**: Investigation. **V.T.**: Investigation. **K.B.**: Investigation. **E.A.**: Investigation. **T.A.**: Investigation. **R.S.T.**: Investigation. **Y.C.**: Investigation. **A.N.**: Investigation. **Y.L.**: Formal Analysis. **N.J.**: Investigation. **R.G.**: Investigation. **T.L.**: Investigation. **J.M.**: Investigation. **S.C.**: Investigation. **M.K.**: Investigation. **J.U.**: Formal Analysis. **T.A.**: Investigation. **J.K.N.**: Formal Analysis. **A.E.**: Investigation. **L.M.**: Investigation. **T.D.**: Investigation. **K.N.L.**: Formal Analysis. **T.S.**: Investigation. **B.T.**: Investigation. **K.W.**: Formal Analysis. **N.S.**: Investigation. **M.M.**: Investigation, Formal Analysis. **A.K.**: Investigation, Formal Analysis. **M.C.**: Investigation, Formal Analysis. **H.G.**: Investigation, Formal Analysis. **K.K.**: Resources. **C.G.**: Consultant, Writing – Review & Editing. **S.K.D.**: Consultant. **C.G.**: Resources. **Y.E.L.**: Resources, Data Curation. **M.W.M.**: Resources, Investigation, Writing – Review & Editing. **K.A.P.**: Resources, Investigation. **A.H.**: Investigation. **J.A.R.**: Resources, Investigation, Writing – Review & Editing. **W.B.**: Investigation, Resources. **P.S.**: Investigation, Resources. **H.C.**: Investigation, Resources. **J.P.**: Investigation, Resources. **C.M.G.:** Investigation, Resources. **Z.D.**: Investigation, Resources. **E.P.**: Investigation, Resources. **C.E.P.**: Consultant, Writing – Review & Editing. **F.K.**: Consultant. **D.A.**: Formal Analysis. **A.I.**: Investigation, Resources. **M.H.**: Investigation. **R.H.**: Investigation. **R.F.**: Investigation. **S.T.**: Consultant, Resources. **L.A**.: Investigation. **X.C.**: Investigation. **R.D.M.**: Investigation, Writing – Review & Editing. **F.S.C.**: Investigation. **J.N.**: Methodology, Validation, Formal Analysis, Investigation, Resources, Data Curation, Writing – Review & Editing, Visualization, Supervision, Project Administration **P.I.D.**: Methodology, Validation, Formal Analysis, Investigation, Resources, Data Curation, Writing – Review & Editing, Visualization, Supervision, Project Administration **J.M.**: Methodology, Validation, Formal Analysis, Investigation, Resources, Data Curation, Writing – Review & Editing, Visualization, Supervision, Project Administration, Funding Acquisition. **T.N.T.**: Conceptualization, Methodology, Software, Validation, Formal Analysis, Investigation, Resources, Data Curation, Writing – Original Draft, Visualization, Supervision, Project Administration, Funding Acquisition.

